# The Influence of Vaginal Microbiome and Clinical Factors on HPV Clearance: A Prospective Study

**DOI:** 10.1101/2022.11.20.22281206

**Authors:** Zhongzhou Yang, Hao Wu, Xiaohan Wang, Guoqing Tong, Zhuoqi Huang, Jie Wang, Yuxin Jiang, Min Cao, Yue Wang, Xingxing Deng, Na Liu, Le Qi, Mengping Liu, Andrew Hutchins, Bin Yao, Mang Shi, Yantao Li, Shida Zhu

## Abstract

**Background:** Although the microbiome and lifestyle factors are associated with HPV clearance, few studies have systematically explored the relevant factors. This formal follow-up prospective study aims to predict HPV clearance based on vaginal microbiota and lifestyle clinical factors.

**Participants and Methods:** Participants were recruited through a digital eHealth platform. Participants were unvaccinated for HPV and were assessed at baseline and a follow-up consultation between August 2021 and January 2022. Both clinical factors and cervicovaginal mucus (CVM) samples were collected from each participant. CVM samples were used to detect HPV and characterize vaginal microbiome by metagenomics. Lifestyle clinical factors were grouped into low-, middle-, and high-risk to operate the stratified analysis as well as survival analysis for HPV clearance.

**Results:** We recruited 141 HPV-positive women at baseline. For the first follow-up, there were 116 HPV persistent infection and 25 HPV clearance participants. Among 28 clinical factors, six factors were identified as significantly associated: age, age of first sexual intercourse, diet balance, marital status, abortion and physical activity under the stratified analysis. Those with a middle-risk diet balance had an odds ratio (OR) (3.91, 95% CI: 1.02-28.03). Those with extremely high-risk diet balance also had a high OR (11.26, 95% CI: 1.35-122.14), but with a faster and higher proportion of HPV clearance. Conversely, clinical factors with low risk and correspondingly faster HPV clearance were physical activity, sleep quality, depression, and anxiety. Although HPV clearance was unrelated to each microbiome specie, HPV clearance was related to the lower overall diversity of species in the vaginal microbiome and the larger abundance of *lactobacillus inners*.

**Conclusions:** This study systematically depicts HPV clearance influenced by clinical factors and its relationship with the vaginal microbial ecosystem. HPV clearance can be improved by modulation of lifestyle habits and marital relationship. The findings from this prospective study have implications for the future design of guidelines to control cervical cancer or other HPV-related cancer, and therefore might be beneficial to women infected with HPV.

## Introduction

Human papillomavirus (HPV) clearance is crucial for reducing the risk of cervical cancer among women [1]. Among men, HPV clearance has also been found to be significantly associated with not having more than one lifetime sexual partner and with increasing age [2]. Oral HPV clearance has been associated with other clinical factors that can reduce the risk of oropharyngeal cancers [3]. This study seeks to systematically explore clinical factors influencing HPV clearance among women. Hence, we conducted this prospective follow-up study to understand the impact of vaginal microbiome and clinical factors for HPV clearance.

In our pilot study, HPV clearance has been proved to be influenced by age, salary, history of reproductive tract infection and total sexual partners. Lifestyle behavior factors are associated with the cancer development and progression [4], for example, physical activity, diet habit, sleep quality score, depression and anxiety scores [5]. However, few studies focused on exploring these lifestyle behavioral factors with HPV clearance. In the vaginal microbiota (VMB) study, it showed that anaerobic VMB was the causative agent of cervical cancer [1] as well as its connection with personal factors [6]. For VMB with the increased abundance of *Lactobacillus spp*, they were more likely related to the HPV clearance [7]. Individual personal features might be beneficial for preventing persistent HPV infection and enhancing HPV clearance through VMB.

Our research explores the relationship between lifestyle behavioral factors and HPV clearance. We explore the associations between long-term HPV infection status (clearance or persistent infection) and VMB through metagenomic sequencing technology as well as HPV genotyping. A digital eHealth platform tool was developed to record age, lifestyle factors, and disease status through standardized questionnaires every three months [8]. In order to reduce heterogeneity, our eHealth platform was developed to collect data among various types of individual lifestyle factors. Using our digital eHealth platform, we systematically collected data on personal factors possibly associated with HPV infection based on existing studies [9]. These factors include: demographics, personal disease history [10], lifestyle behavior on malnutrition [11], sexual history [12-14] and number of sexual partners, and substance abuse (especially smoking habits) [15].

## Methods

### Study design and samples

The research ethics of this prospective study was obtained from the Institutional Review Board of the Beijing Genomics Institution (BGI-IRB 21078). The study was also recorded in the Chinese clinical trial registry (ChiCTR) website (www.chictr.org.cn, ChiCTR2100049221). The Ministry of Health designed the ChiCTR to present Chinses clinical study in the World Health Organization International Clinical Trial Registration Platform (WHO ICTRP). It provides the platform of study design and research information available to the public and global sharing in WHO ICTRP.

The inclusion criteria were nonpregnant and nonlactating women for eligible participants. They were also unvaccinated for HPV but had sexual experience at least once in their lifetime and had a diagnosed HPV infection in the last 12 months. All eligible patients wrote the informed consent to authorize the research team to utilize their anonymized eHealth personal data of clinical personal factors. After successfully enrolling in this trial, the baseline recruitment of participants for this study took place from 25 August to 24 September 2021 and the baseline HPV infection was recorded. For the first follow-up, participants were recruited from 25 October to 24 November 2021. Based on their follow-up HPV test results, participants were grouped into two cohorts: HPV-clearance defining as the baseline HPV positive conversion to first follow-up HPV negative. Then, HPV persistent infection was defined as HPV-positive for both baseline and follow-up diagnosis. Samples were collected same as the pilot study described elsewhere [16].

### Laboratory test and eHealth Platform

The samples operated for both HPV test and metagenomics. Detailed protocol was shown in our pilot study [16]. The basic function of eHealth platform and personal record outcomes (PROs) were described in the pilot study [16]. PROs covered seven categories as demographic information, medical history, lifestyle factors concerning physical activity, diet balance, sleep quality, depression, and anxiety (S3-S6 questionnaire content), sexual history and behavior, and substance abuse. The lifestyle behavior scores were grouped same with our HPV infection study [17]

### Quality control and taxonomic annotation

The sequencing reads were quality-controlled by fast-protein (fastp) [17]. Front bases with a quality score of less than 20 were cut and reads with lengths shorter than 30 bp were removed. Host contamination reads were removed using bowtie2 [18] and hg38 index parameters *‘*--very-sensitive*’*. Unaligned reads were retained for analysis. Taxonomic assignment of high-quality metagenomic shotgun data was performed using MetaPhlAn2 [19] and functional profiling was performed using HMP Unified Metabolic Analysis Network (HUMAnN 2.0) [20]. The VAginaL community state typE Nearest CentroId clAssifier (VALENCIA) was used for the nearest centroid classification method [21], for community state type (CST) classification. We divided samples into 5 types based on how each was dominated by a different species: for CST I—*Lactobacillus. crispatus* dominated, for CST II—*Lactobacillus. Gasseri* dominated, for CST III—*Lactobacillus. iners* dominated, for CST V—*Lactobacillus. jensenii* dominated, and CST IV was dominated by a species that did not belong to *Lactobacillus*.

### Statistical Analysis

The statistical analysis includes three sections: preliminary demographic analysis of VMB and clinical lifestyle factors, logistic regression for the stratified analysis, and survival analysis. The preliminary demographic analysis was sourced from our pilot study [16]. The logistic regression was conducted same with HPV infection study for HPV clearance and lifestyle factors [17]. Survival analysis was conducted for lifestyle factors and community state type. It required the survival time, outcome category, and scores. To determine the period from baseline to first follow-up, we extracted the associated delivery date of baseline data and first follow-up for HPV infection as well as HPV clearance. Then the survival time was defined as the period from baseline to first follow-up for each participant.

Categories depended on whether participants had an HPV infection or clearance outcome during their first follow-up. Furthermore, we calculated alpha diversity and bray distance based on R version (4.0.4). In addition, species and pathways were filtered for occurrence rates of less than 10%. A Wilcoxon test was performed to explore the changes in vaginal microbiome between HPV persistent infection and HPV-clearance individuals. The Kruskal-Wallis test was used to explore the relationship between vaginal microbiome and lifestyle clinical factors.

## Results

### Participant Recruitment and HPV Cohort Grouping

In our study, 141 participants were recruited in the baseline period through digital eHealth tool. After three months follow-up, we identified 116 HPV-persistent and 25 HPV-clearance participants. Table 1 showed the patient features for HPV infection. The average age was 38.11 (SD: 10.30) for HPV-persistent infection participants and the average age for HPV-clearance participants was 42.46 (SD: 8.17), which was significantly different (P-value = 0.029). For educational history, occupation, and salary, these factors were insignificant between HPV infection and clearance. Disease status was not statistically related to HPV infection, denoting our recruitment was not biased because of other clinical diseases. For sexual activity realted factors, those that were significant included marital status (P-value = 0.0499), abortion (P-value = 0.02193) and age of first sexual intercourse (P-value = 0.0455). However, other clinical variables were insignificant, such as smoking, alcohol habits, and hospital detection.

**Table 1.**
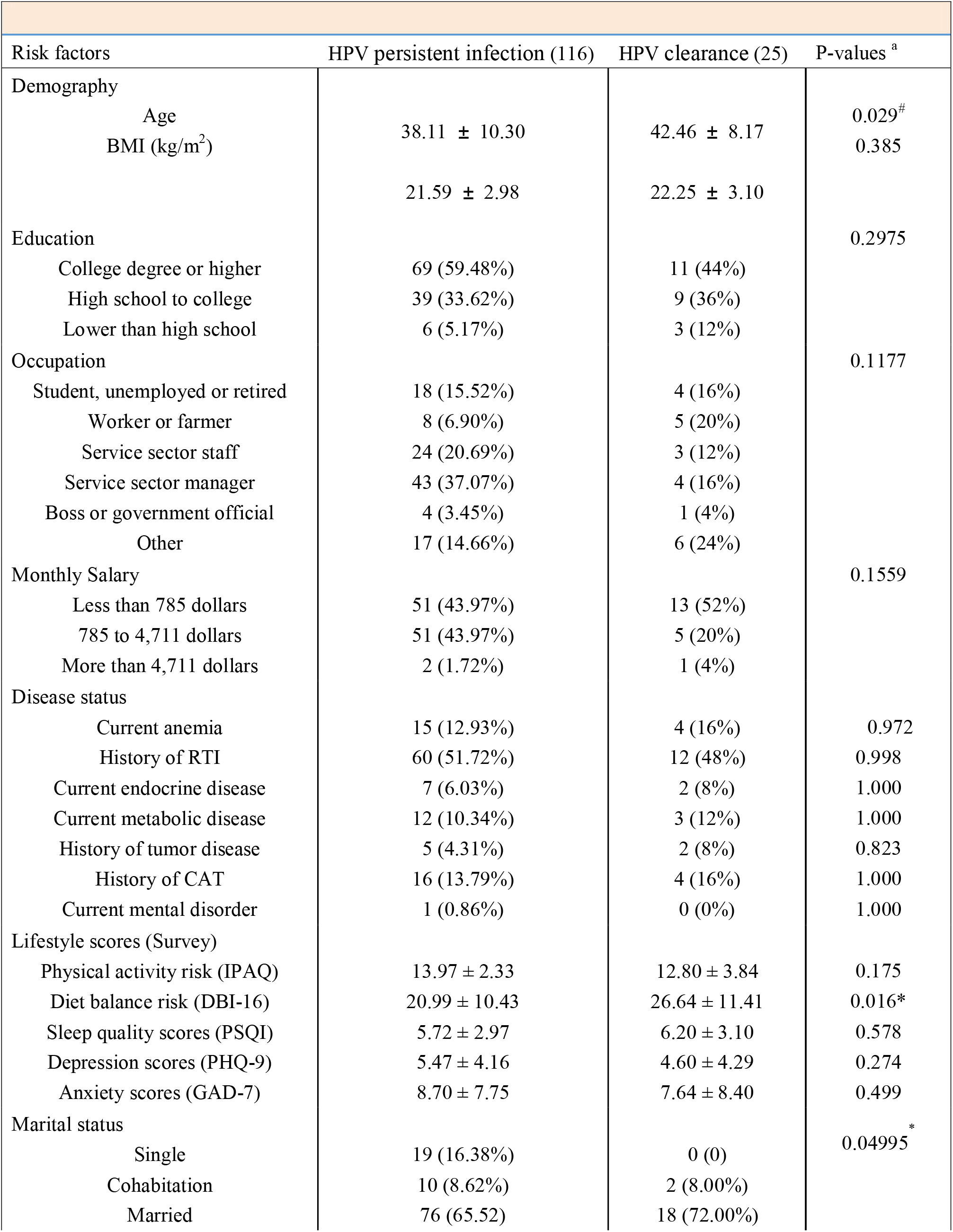
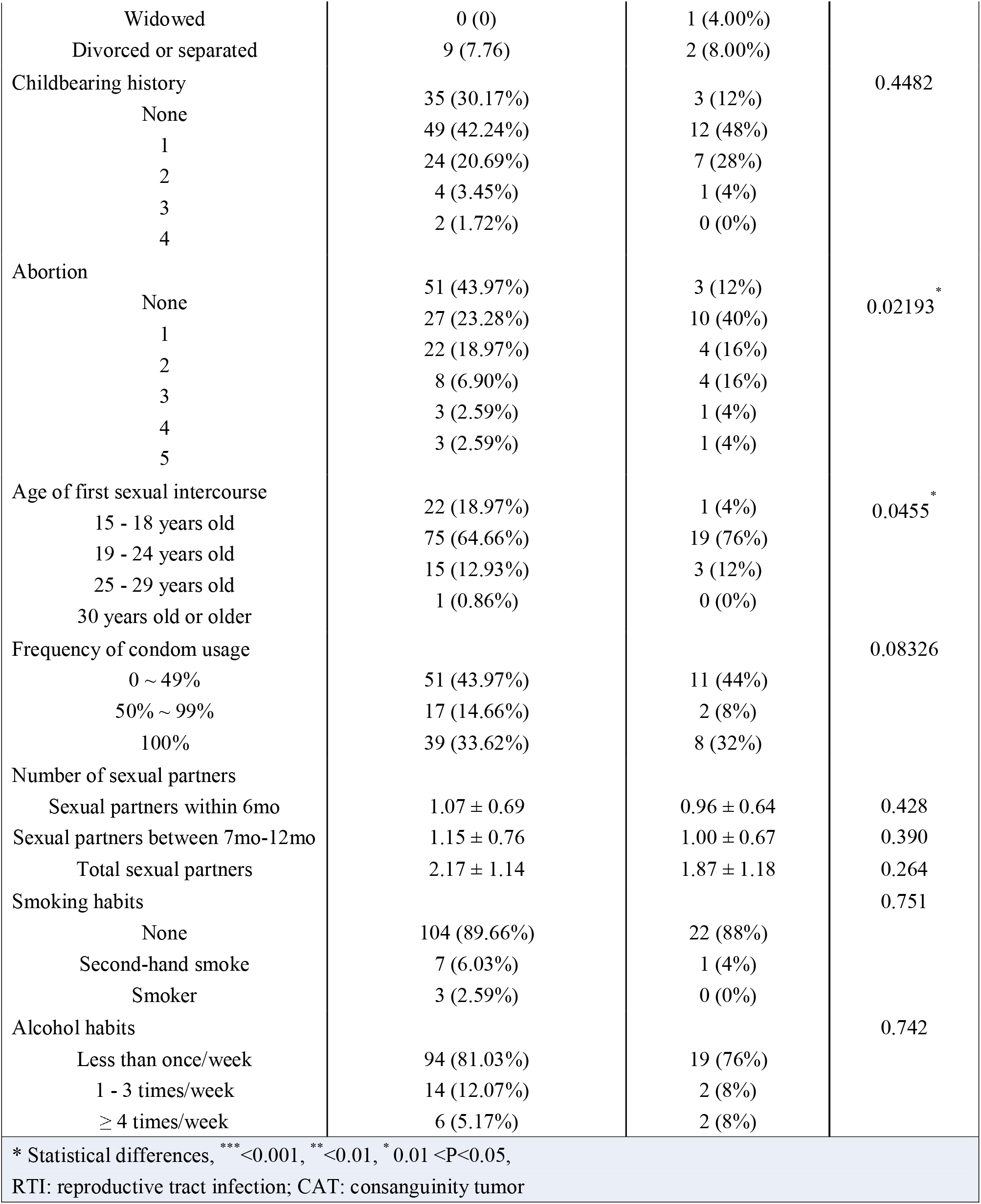
Characteristics of 141 participants included in study design

In Table 1, we also considered the clinical lifestyle factors and vaginal microbiome community state type (CST). Among lifestyle behavior scores, surprisingly, diet balance was statistically significant (P-value = 0.016). However, the other four lifestyle factors, physical activity, sleep quality, depression, and anxiety scores were not significant. A lower diet balance score was significantly correlated with a higher risk of HPV infection, when comparing the HPV-persistent infection group (20.99) versus the HPV-clearance group (26.64). In terms of vaginal microbiome population makeup, it was statistically significant (P-value = 0.002) when combining CST I ∼ V. The complete survey is shown with a range of 76% ∼ 100% in Supplementary Table 1. The data that support the findings of this study have been deposited into CNGB Sequence Archive (CNSA) [22] of China National GeneBank DataBase (CNGBdb) [23] with accession number CNP0003307.

### Vaginal Microbiome

In our data, 223 species from 105 genera were identified to perform CST typing as well as PCOA. Changes between baseline and follow-up samples in our analysis showed that CST III is an intermediary type. The switch always occurred between CST I and CST II, which we regard as the healthier type, or between the unhealthy type CST IV and CST III. We hardly ever found direct jumps for I and IV. (Supplementary Figure 1)

We calculated alpha diversity at the species level for each sample and found that the diversity of HPV-persistent infection samples was significantly higher than HPV clearance samples (P-value=0.049) (Figure 1A). In addition, after comparing 25 baseline and follow-up individual samples with HPV-clearance, we found that diversity tends to decrease (P-value=0.07) among these participants (Supplementary Figure 2).

**Figure 1.**
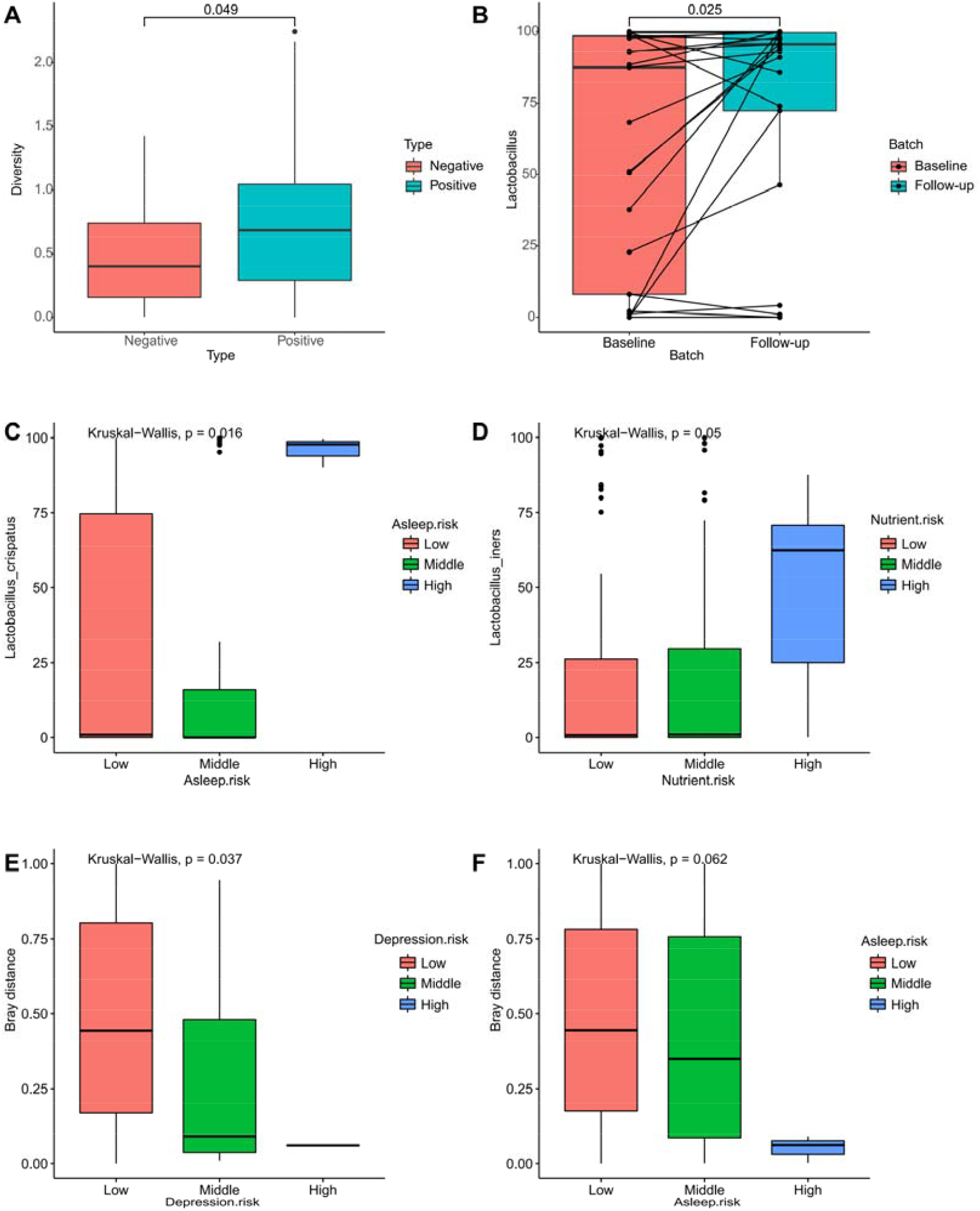
Taxonomy difference analysis between groups. Legend: A. Alpha-diversity represented in HPV-persistent infection and HPV-clearance samples indicating significant difference (P-value = 0.049) based on the Wilcoxon test. The boxes represent distributions of the alpha diversity index. B. The abundance of Lactobacillus represented in HPV-clearance samples indicating significant difference (P-value = 0.025) between baseline and follow-up groups based on pairwise Wilcoxon test. The boxes represent distributions of the abundance of *Lactobacillus*. Lined samples are from the same individual. Some species and bray distances show difference under different personal factor risk levels based on the Kruskal-Wallis test. (C) *L*.*crispatus* with asleep risk level, (D) *L*.*iners* with Nutrient risk level, (E) bray distance with depression risk level, and (F) bray distance with asleep risk level.

Furthermore, genus-level annotation showed a significant increase of *Lactobacillus* (P-value=0.025) (Figure 1B). Although the species level did not show any significant difference, the bacteria showed a statistically significant correlation (P-value<0.1) related to CST typing (Supplementary Table 2).

Meanwhile, we further analyzed the dominant bacteria associated with CST typing. The metagenomic dataset is shown as seventeen items in Table 2. For lactobacillus-related factors, *Lactobacillus crispatus* had the largest proportion of serotype for HPV infection at 32.3 (SD: 42.8). However, the largest proportion of HPV clearance belonged to lactobacillus iners at 38.6 (SD: 42.7). Nonetheless, both crispatus and iners were insignificant serotypes, as well as other lactobacillus-related serotypes. For gynecological disease related to serotype, harmful *Gardnerella vaginae* had the largest proportion for both HPV infection and clearance. Moreover, *Atopobium vaginae* was the pseudo-significant serotype (P-values = 0.057) and other harmful serotypes were also insignificant between HPV infection and clearance.

**Table 2.** Relative abundance of bacteria

| Table 2. Relative abundance of bacteria |  |  |  |
| --- | --- | --- | --- |
| Risk factors | HPV persistent infection (116) | HPV clearance (25) | p-Values <sup>a</sup> |
| Lactobacillus-related (6) |  |  |  |
| Crispatus | 32.3 ± 42.8 | 21.0 ± 37.4 | 0.409 |
| Gasseri | 0.1 ± 0.7 |  | 0.195 |
| Iners | 16.7 ± 27.4 | 0.2 ± 0.8 | 0.071 |
| Jensenii | 1.3 ± 4.6 |  | 0.898 |
| Lactobacillus_other | 8.0 ± 21.8 | 38.6 ± 42.7 | 0.943 |
| Other_microbes |  |  | 0.176 |
|  | 19.2 ± 30.4 | 7.2 ± 21.1 |  |
|  |  | 7.0 ± 20.4 |  |
|  |  | 11.5 ± 24.0 |  |
| Harmful-related (11) |  |  |  |
| Trachoma_chlamydia |  | 0 ± 0 |  |
| Neisseria_gonorrhoeae |  | 0 ± 0 |  |
| Microureaplasma |  |  |  |
| Mycoplasma_hominis |  | 0.4 ± 0.8 |  |
| Candida_albicans |  |  |  |
| Gardnerella_vaginae | 0 ± 0.2 |  | 0.353 |
| Prevotella_bivia | 0 ± 0 | 0.2 ± 1.0 | NA |
| Atopobium_vaginae | 0.4 ± 1.4 |  | 0.472 |
| Diallisteria | 0.3 ± 1.8 | 0 ± 0 | 0.958 |
| Streptococcus_agalactiae | 0 ± 0 |  | NA |
| Timona_prevotella | 19.1 ± 29.8 | 12.9 ± 25.5 | 0.094 |
|  | 2.4 ± 9.9 |  | 0.093 |
|  | 3.3 ± 10.5 | 0.3 ± 1.0 | 0.057 |
|  | 0 ± 0.1 |  | 0.307 |
|  | 0 ± 0 | 1.7 ± 6.0 | 0.518 |
|  | 0.5 ± 2 |  | 0.426 |
|  |  | 0 ± 0 |  |
|  |  | 0 ± 0 |  |
|  |  | 0.1 ± 0.4 |  |
| <sup>a</sup> Statistical difference from two-sample Wilcoxon tests |  |  |  |

**Table 3.** Stratified analysis

| Table 3. Stratified analysis |  |  |  |  |  |
| --- | --- | --- | --- | --- | --- |
| Risk factors | HPV persistent infection (116) | HPV-clearance (25) | Odds ratio | 95% CI | P-values |
| Physical activity |  |  |  |  |  |
| High level | 3 | 4 | 1.00 | NA | NA |
| Moderate level | 18 | 3 | 7.10 | 1.03 - 60.44 | 0.02<sup>#</sup> |
| Low level | 95 | 18 | 6.79 | 1.32-39.58 | <0.01<sup>##</sup> |
| Diet balance risk |  |  |  |  |  |
| Almost no problem | 3 | 2 | 1.00 | NA | NA |
| Low level of under intake | 81 | 17 | 3.20 | 0.35-22.56 | 0.27 |
| Moderate level of under intake | 30 | 6 | 3.26 | 0.32-26.32 | 0.29 |
| High level of under intake | 2 | 0 | NA | NA | NA |
| Sleep quality risk |  |  |  |  |  |
| Good | 87 | 18 | 1.00 | NA | NA |
| Moderate | 26 | 7 | 1.31 | 0.46-3.41 | 0.60 |
| Bad | 3 | 0 | NA | NA | NA |
| Depression risk |  |  |  |  |  |
| Minimal | 49 | 13 | 1.00 | NA | NA |
| Mild | 56 | 8 | 1.84 | 0.71-5.06 | 0.21 |
| Moderate | 6 | 4 | 0.4 | 0.10-1.84 | 0.23 |
| Moderately severe | 3 | 0 | NA | NA | NA |
| Severe depression | 2 | 0 | NA | NA | NA |
| Anxiety risk |  |  |  |  |  |
| Minimal | 49 | 13 | 1.00 | NA | NA |
| Mild | 15 | 4 | 0.98 | 0.29 - 4.01 | 0.97 |
| Moderate | 12 | 0 | NA | NA | NA |
| Severe | 40 | 8 | 1.32 | 0.50-3.67 | 0.58 |
| IPAQ = international physical activity questionnaires. DBI = diet balance index. PSQI = Pittsburgh sleep quality index. PHQ-9 = Patient Depression Questionnaire-9. GAD 7 = Generalized Anxiety Disorder 7. NA = Not available |  |  |  |  |  |

### Logistic Regression for Lifestyle Factors

To evaluate the association between lifestyle factors and HPV clearance, univariate logistic regression was performed for five lifestyle behavior factors (Table 2). Among the five factors, physical activity significantly increased middle-risk by 7.10 (95% CI: 1.03 - 60.44, P-value = 0.02) and high-risk by 6.79 (95% CI: 1.32-39.58, P-value < 0.01). For diet balance scores, middle-risk significantly increased the odds ratio by 3.91 (95% CI: 1.02-28.03, P-value < 0.01) and extremely high-risk increased the odds ratio by 11.26 (95% CI: 1.35-122.14, P-value < 0.01). For the other three lifestyle factors, there were three odds ratios unavailable including those for high-risk sleep quality risk, high-risk depression risk and middle-risk anxiety risk, because of zero participants in the corresponding categories.

### Correlation between Personal and lifestyle Factors and Microbiome

After excluding species with an occurrence rate of less than 10%, correlation analysis showed that *L*.*crispatus* had significant difference in each sleep quality status (P-value=0.016) (Figure 1C), and *L*.*iners* correlates with diet balance (P-value=0.05) (Figure 1D). Furthermore, the bray distance implies a higher the risk of depression (P-value=0.037) (Figure 1E) and worse sleep scores (P-value=0.062) between baseline and follow-up samples (Figure 1F).

### Survival Analysis

To predict the clearance rate and period, both lifestyle factors and CST were assessed using survival analysis (Figure 2). Figure 2A shows low-risk physical activity (57%) with the largest clearance ratio compared to middle-(18%) and high-risk (16%). However, diet balance shows the inverse pattern with physical activity (Figure 2B). Moreover, physical activity had a consistent correlation with sleep quality (Figure 2C), depression (Figure 2D) and anxiety (Figure 2E). For CST survival analysis in Figure 2F, CST III was more likely to achieve clearance at 26%, but both CST II and V were unlikely to achieve HPV clearance (0%).

**Figure 2:**
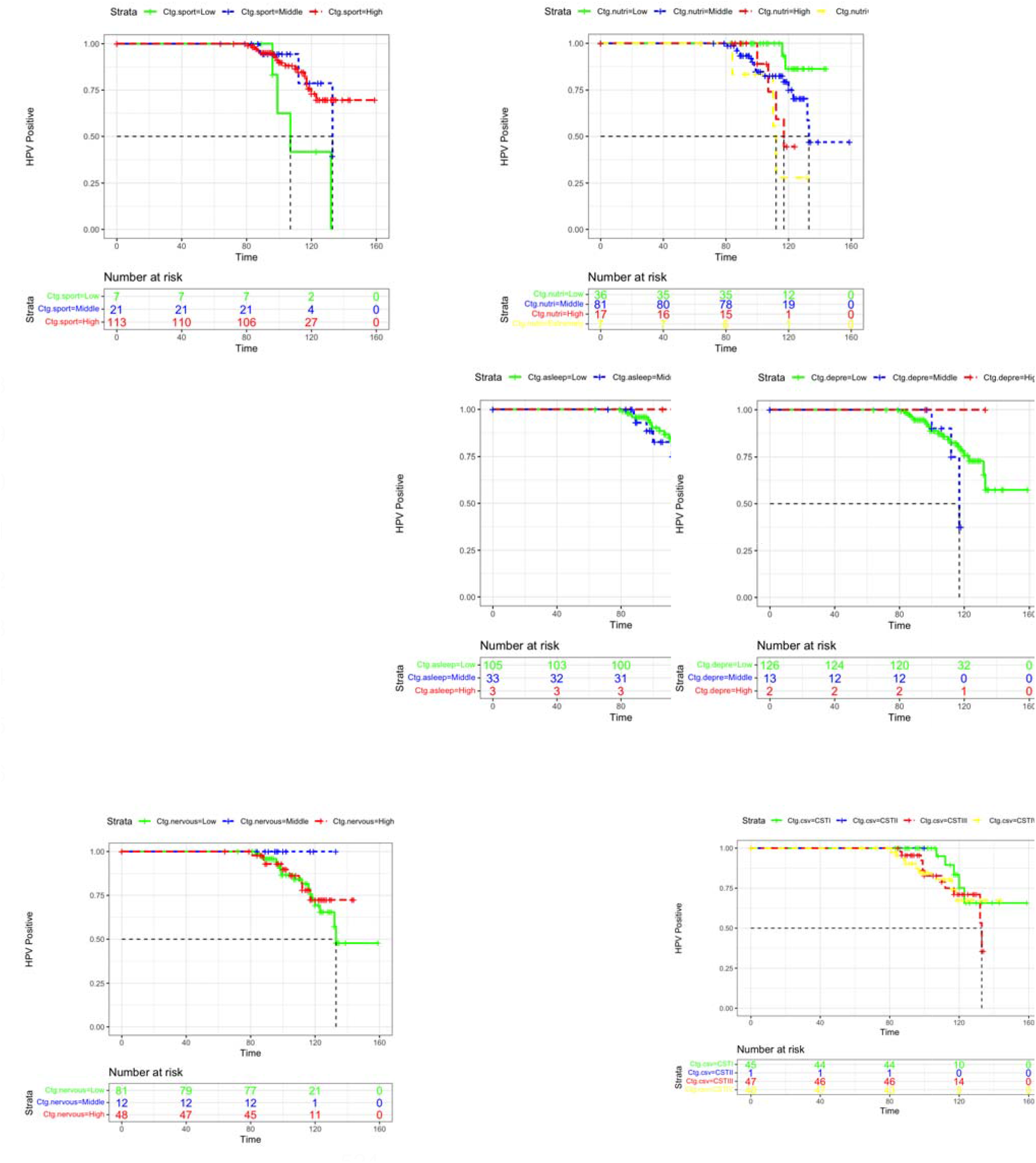
Lifestyle factors for survival analysis. Legend: survival analysis for lifestyle clinical factors among (A) Physical activity, (B) Diet balance, (C) Asleep quality scores, (D) Depression scores, (E) Nervous scores; Color coding: green = low risk, blue = middle risk, red = high risk, yellow = extremely high risk for diet balance. Survival analysis for (F) Community state type. Color coding: green = CST I, blue = CST II, red = CST III, yellow = CST IV

At the functional level, 347 pathways were annotated and 66% of them had an occurrence rate of more than 5%. Coenzyme A synthesis was highly relevant for two pathways including PWY-4242 (P-value=0.022) and COA-PWY (P-value=0.044), with significant difference between negative and positive samples. Individual HPV-clearance was significantly increased with, an abundance of peptidoglycan synthesis pathways PEPTIDOGLYCANSYN-PWY (P-value=0.034) (Figure 3). In addition, we also found that PWY.7111 and VALSYN.PWY were significantly changed in two samples of individuals with HPV clearance (P-value=0.025).

**Figure 3.**
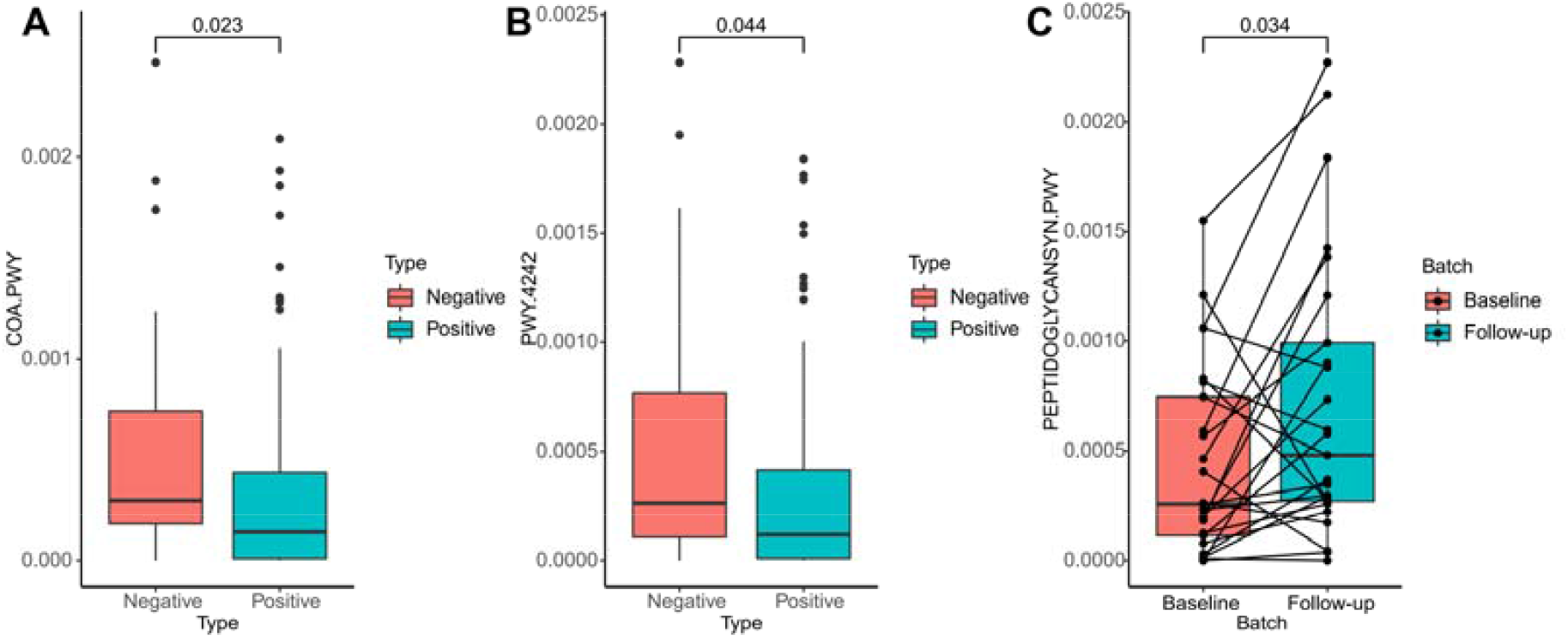
Functional difference analysis between groups. Legend: The abundance of some function pathways indicating significant difference based on Wilcoxon test. The boxes represent distributions of function pathway abundance. Lined samples are from the same individual. (A) COA-PWY between HPV clearance and HPV persistent infection in follow-up samples, (B) PWY-4242 between HPV clearance and HPV persistent infection in follow-up samples, (C) Peptidoglycansyn-pwy between baseline and follow-up in HPV-clearance samples.

## Discussion

A typical limitation of study design exploring clinical lifestyle factors or vaginal microbiota related to HPV clearance, is in defect of prospective studies to evaluate the influence of the lifestyle factors or microbiome on HPV clearance. In this research, we explored the association between clinical lifestyle factors or microbiome and HPV clearance in the prospective cohort of 141 women. Our results suggest that the lifestyle factor of the diet balance at the baseline of HPV infection may significant influence the HPV clearance. Meanwhile, it had the similar findings of current disease or disease history as well as univariate logistic regression for our previous HPV infection study (17).

We found that community state type (CST) might guide the way as the significant indicator for HPV clearance. First, CST III was found to be an intermediary type suggesting that interventions targeting CST III may be more efficient for clearing HPV [7, 24]. Meanwhile, in HPV clearance samples we observed a decrease in biodiversity and increase of *Lactobacillus*. This is consistent with previous studies which found that individuals were healthier when their samples were *Lactobacillus*-dominated and had low diversity. This suggests that enhancing *Lactobacillus* by taking probiotics might be useful for HPV clearance. We also found that a variety of *Lactobacillus* were associated with personal lifestyle factors, suggesting that good lifestyle habits may be crucial for healthier vaginal microbiome, thus promoting HPV clearance.

Coenzyme A (CoA) is the crucial co-factor of cellular metabolism in human beings. Previous studies have shown that the CoA synthesis pathway is one of the important sources of human gut microbes. In this study, we found that the synthesis pathway of CoA was also common in the vaginal microbiome. One research showed that the increased levels of CoA was able to kill cancer cells including cervical and breast cancers [25]. We found that samples with HPV-clearance had a higher abundance of CoA synthesis pathways. This suggests that CoA might also have an inhibitory effect on the process of HPV infection leading to carcinogenesis.

Previous studies have indicated that an abundance of peptidoglycan biosynthesis pathways is more frequent in normal microbiota compared to bacterial vaginosis (BV) samples [26]. We know that infection with BV has a harmful impact on HPV clearance. This suggests that we can utilize peptidoglycan against BV, thereby promoting protection for the HPV clearance process. Above all, the discovery of significantly different pathways suggests the possibility of intervention using coenzyme A and peptidoglycan at the functional level.

Our statistical analysis demonstrated the significant clinical lifestyle factors. To enhance HPV clearance, the stratification analysis reported how the low-risk physical activity was beneficial to HPV clearance. This might be elucidated by the effect of human immune system, such as cell immunological function via CD4/CD8 T cells [27]. Regarding HPV clearance and diet balance, stratification analysis reported the higher diet balance risk leading to the more difficult for clearing the baseline HPV infection (OR: 3.27). The potential reason was high-risk diet balance relating to the lower levels of lycopene or vitamin A for HPV infection [28, 29] in the baseline through the cell-based immune system [30]. Based on our questionnaires, one solution we recommend for improving diet balance is to intake appropriate consumption of vitamin A including vegetables (e.g. tomatoes), dairy products and fruits, in order to attain a low-risk diet balance [31].

After adjusting for physical activity and diet balance, another three lifestyle factors including sleep quality, depression and anxiety showed insignificant related to HPV clearance through logistic regression. In addition, survival analysis provided crucial indicators to predict HPV clearance and approaches to promote clearance under lifestyle risk factors. Those with a low-risk lifestyle for physical activity, sleep quality, depression and anxiety had a higher possibility of clearing HPV. Although those with an extremely high-risk diet balance had a higher possibility of HPV clearance compared to low-risk, the potential reason might be explained by reduced sexual activity for those under extremely high-risk diet balance.

### Strengths

This study has three major strengths. The first strength is establishing the digital eHealth platform enabled participants to fill in their health-related information through mobile phone to decrease the man-made error. The second is systematically collecting lifestyle clinical and sexual factors. Previous studies have only focused on sexual factors and few studies have considered lifestyle factors more broadly. The third is self-sampling for cervical-vaginal mucus, which is a relatively novel feature for gynecological screening. This might help remove personal barriers that hinder women from enrolling in cervical screening programs. Moreover, self-sampling promotes the accessibility of public health interventions by removing the need for a clinical specialist’s physical presence, thereby enabling those even in remote areas to receive screening.

### Weaknesses

Although we have found some obvious trends and results, our study still has some limitations such as an unbalanced sample size and a limited number of negative samples. We still need to follow-up for a longer duration and collect more samples. If there were a test population with a more balanced sample size (especially with more negative samples) for comparison, the corresponding results would have greater statistical power.

### Conclusions

In this study, we systematically depict HPV clearance by clinical factors, specifically lifestyle clinical factors as well as the vaginal micro-ecosystem. HPV clearance can be improved by superior lifestyle habits including physical activity in addition to other established factors relating to sexual life and marital relationship. Our findings might be beneficial to women at higher risk of contracting HPV and provide relevant information for future designs of guidelines for controlling cervical cancer or other HPV-related cancer.

## Author Contributions

Zhongzhou Yang, Yantao Li and Shida Zhu designed the project, analyzed the clinical data, prepared the manuscript, and carried out this study. Hao Wu, Min Cao and Yue Wang conducted laboratory work of HPV test and metagenomics. Xiaohan Wang and Yuxiang Lin analyzed the laboratory data and explored the association between metagenomics and the clinical dataset. Zhuoqi Huang collected the dataset and helped to obtain approval from the relevant ethics committees. Xingxing Deng and Jie Wang were responsible for preparing the sampling kits. Mengping Liu helped to revise the written content. Shida Zhu, Na Liu and Le Qi funded this project. Guoqing Tong and Andrew Hutchins helped to further improve the quality of the manuscript.

## Data Availability

All data produced in the present study are available upon reasonable request to the authors

## Acknowledgements

We acknowledge the funding source of Shenzhen Engineering Laboratory for Innovative Molecular Diagnostics (DRC-SZ [2016]884). Meanwhile, we also acknowledge BGI IT team (Chungen Zhang and Weiwei Bai) for developing the eHealth platform and BGE team (Kaiye Cai). Finally, we acknowledge Tim Yung for language revision.

## Figure Legends

**Supplementary Figure 1.**
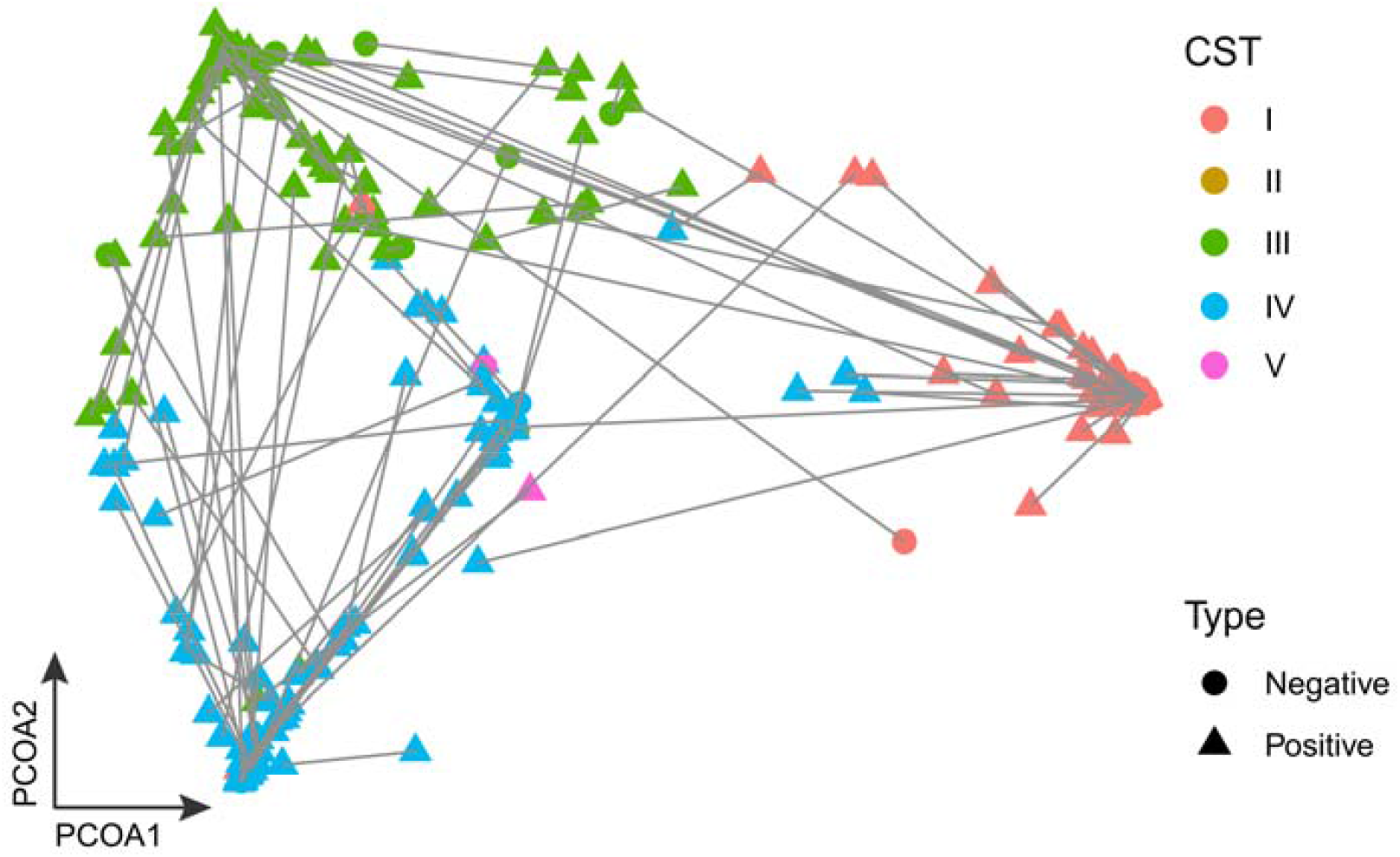
Legend: PCoA distribution of all samples. Different colors represent different CST types, and different shapes represent HPV clearance and HPV persistent infection. Lined samples are from the same individual.

**Supplementary Figure 2.**
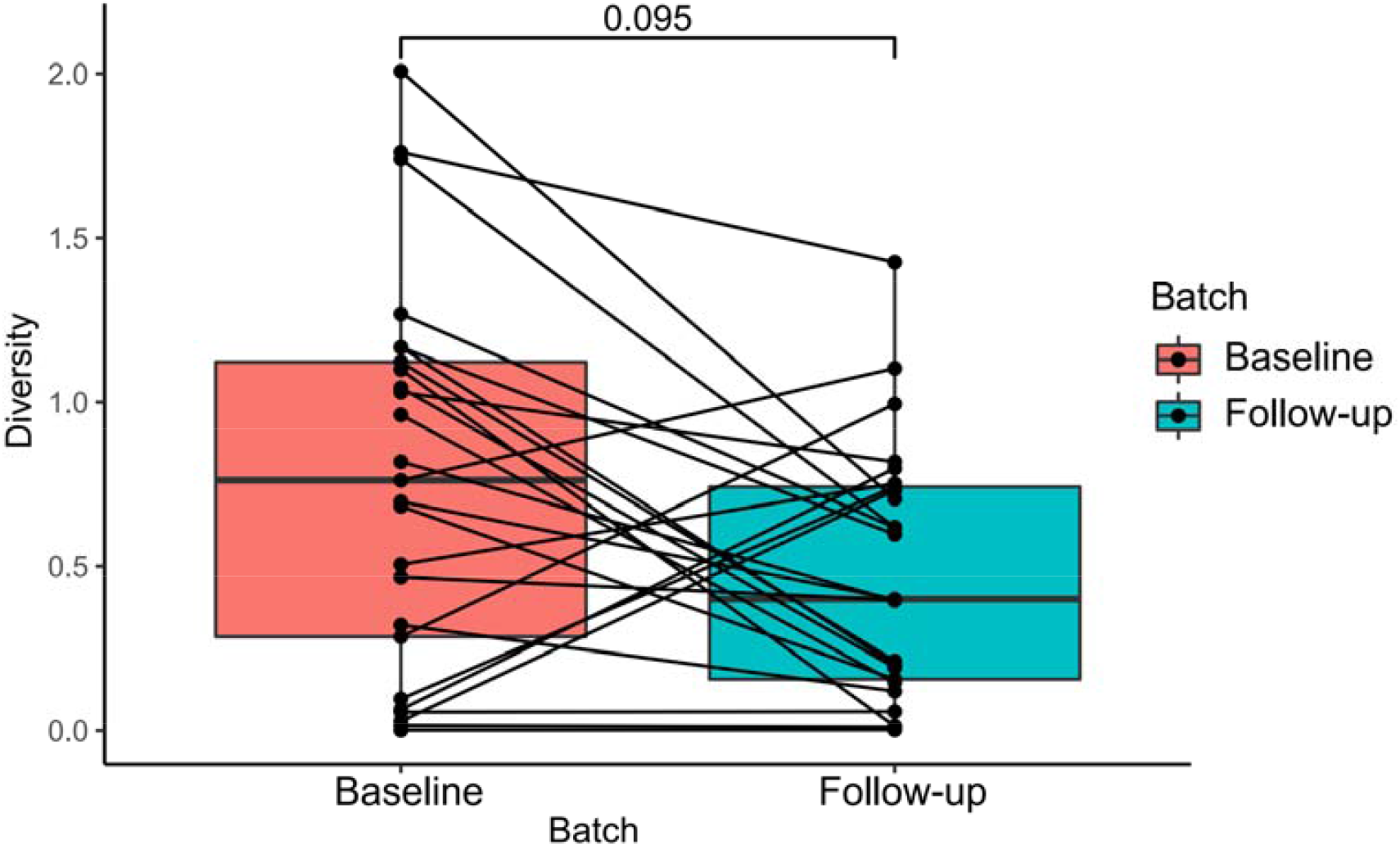
Legend: Alpha-diversity represented in HPV clearance samples indicating difference (P-value = 0.095) between baseline and follow-up groups based on Wilcoxon test. The boxes represent distributions of the alpha diversity index. Lined samples are from the same individual.

## Notes

### Competing Interest Statement

The authors have declared no competing interest.

### Funding Statement

This study did not receive any funding

### Author Declarations

Ethics committee/IRB of Beijing Genomics Institution (BGI-IRB 21078) gave ethical approval for this work.

